# Symptom-based testing in a compartmental model of COVID-19

**DOI:** 10.1101/2020.10.11.20211037

**Authors:** Ferenc A. Bartha, János Karsai, Tamás Tekeli, Gergely Röst

## Abstract

Testing and isolation of cases is an important component of our strategies to fight SARS-CoV-2. In this work, we consider a compartmental model for COVID-19 including a nonlinear term representing symptom-based testing. We analyze how the considered clinical spectrum of symptoms and the testing rate affect the outcome and the severity of the outbreak.

## 1 Introduction

Since a cluster of pneumonia cases of unknown origin was discovered in Wuhan, China in late 2019, COVID-19, the disease caused by the novel coronavirus SARS-CoV-2, has spread around the world giving rise to a pandemic. By early August 2020, around eighteen million cases and seven hundred thousand deaths have been reported worldwide [1].

One of the key difficulties in controlling COVID-19 is that many infections result in mild symptoms or none at all, making the detection of infectious COVID-19 cases particularly challenging [2]. Moreover, a considerable portion of secondary infections, generated by those infectors who later develop symptoms, have been observed to take place before symptom onset, *i.e*. pre-symptomatic transmission occurs [3, 4, 5].

COVID-19 is classified as a respiratory disease, accordingly, it mainly affects the respiratory tract (similarly to other coronaviruses) but other classes of symptoms have been observed as well, *e.g*. affecting the gastrointestinal [6] and musculoskeletal systems [7]. In particular, the loss of smell (anosmia) and/or taste (ageusia) could be a key indicator symptom of COVID-19 [8]. We summarize findings of some recent studies regarding COVID-19 symptoms and their prevalence among clinical COVID-19 cases in Table 1.

**Table 1:**
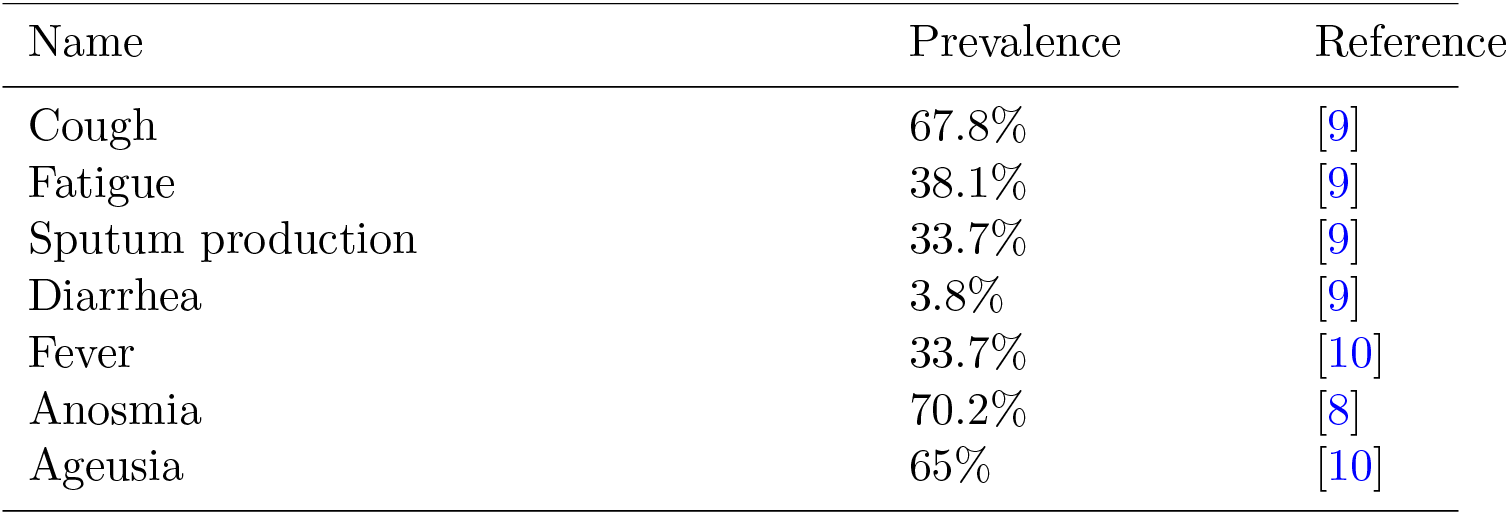
Several key symptoms of COVID-19 and their prevalence among <u>clinical COVID-19 cases.</u>

| Name | Prevalence | Reference |
| --- | --- | --- |
| Cough | 67.8% | [9] |
| Fatigue | 38.1% | [9] |
| Sputum production | 33.7% | [9] |
| Diarrhea | 3.8% | [9] |
| Fever | 33.7% | [10] |
| Anosmia | 70.2% | [8] |
| Ageusia | 65% | [10] |

The primary confirmation of COVID-19 infection, as of now, happens via real-time reverse transcription polymerase chain reaction (rRT-PCR) based testing of samples taken from *e.g*. nasopharyngeal or oropharyngeal swabs, sputum, lower respiratory tract aspirates, etc [11].

A classical approach for modeling and understanding epidemics is constructing a system of ordinary differential equations (ODE) having a compartmental structure. Also, such models are widely utilized as an important tool of assessing the effectiveness of various control strategies [13, 14].

In particular, the transmission dynamics of the spread of COVID-19 has been analyzed via compartmental ODE models in a vast number of studies. Yang and Wang [15] investigated the outbreak of COVID-19 in Wuhan, China considering multiple transmission pathways in the infection dynamics. Non-constant transmission rates were employed, changing with the epidemiological status and environmental conditions reflecting the impact of the ongoing disease control measures. Boldog et al. [16] developed a tool comprised of three major components to assess the risk of global spread of COVID-19 with origin from Wuhan. A time-dependent *SEIR* model (Suceptible-Exposed-Infectious-Removed) was used to estimate the cumulative number of cases in China from which probability distributions were obtained for the number of potential disease spreaders outside China. Finally, for a given destination country, the initial spread of COVID-19 was approximated via a Galton-Watson process. Berger et al. [17] utilized an extended *SEIR* model to understand the role of testing and case-dependent quarantine with fixed rates and compared simple testing and quarantine policies. Weitz [18] developed an extended, age-stratified model analyzing both asymptomatic and severe courses of the disease in order to estimate the burden on the healthcare system by modeling hospital and intensive care unit (ICU) bed needs. Röst et al. [19] studied an age-stratified compartmental model and presented a comprehensive analysis exploring several post-lockdown scenarios with age-specific measures, seasonality, and spatial heterogeneity.

In this work, we consider an extended *SEIR*-type compartmental model for the transmission dynamics of COVID-19. We incorporate symptom-based testing of patients and isolation upon positive result *i.e*. removal from the infectious chain. The clinical symptoms that trigger the testing of individuals is referred to as *indicator symptom*. The *force of testing* is defined as the rate at which infected individuals are tested, see Sect. 2. It is described by a nonlinear function of the state of the epidemic and of all individuals displaying the indicator symptom at a given time, with or without COVID-19 infection, hence, it is considerably different from previous approaches. Our goal is to understand the impact, and especially the limitations of this testing strategy, hence we model neither contact-tracing of patients with positive tests nor the testing of a fraction of non-symptomatic contacts, both of which are common and efficient improvements and result in removal of additional patients from the infectious chain. Moreover, we assume perfect testing, that is we do not consider false positive or false negative results.

According to the current understanding of the disease, none of the symptoms are specific solely for COVID-19, thus, the chosen indicator symptom may and will be present amongst other individuals not infected with SARSCoV-2. All patients, with or without COVID-19 infection, displaying the indicator symptom form the so-called *primary symptom pool*, whilst, those without COVID-19 infection (but with the same indicator symptom) are members of the *secondary symptom pool*, see Sect. 2. Naturally, choosing the indicator symptom for a testing campaign should be affected by its prevalence and by the historical statistics for the size of the associated secondary symptom pool. We emphasize that the latter might undergo seasonal variations as is typical with respiratory symptoms peaking in influenza season [20]. This is a common but not uniform feature of COVID-19 symptoms, *e.g*. gastrointestinal symptoms might show no seasonal variations, depending on age-groups [21].

The chapter is structured as follows. Sect. 2 presents the compartmental epidemic model and its parametrization. In addition, the next generation matrix computations are included that are used to derive formulae for the reproduction number. Then, Sect. 3 establishes several boundedness and monotonicity-type results on key characteristics of the epidemic model. The results of numerical simulations are discussed in Sect. 4. Finally, we present our conclusions in Sect. 5.

## 2 The epidemic model of indicator symptom-based testing

To assess the effectiveness of indicator symptom based testing in controlling the spread of COVID-19, we developed a compartmental population model based on the general *SEIR* formulation without vital dynamics.

We divide the population into five classes: susceptible (*S*), latent (*L*), pre-symptomatic (*P*), infected (*I*), and removed (*R*). Susceptibles are those who can get infected by SARS-CoV-2. The members of the latent compartment *L* have already been infected, but are not yet infectious nor do they display any symptoms. After that, latent individuals move to the pre-symptomatic class *P* meaning that, due to the increased viral load, they are able to infect susceptible individuals, even though, they still not display any symptoms. The existence of pre-symptomatic transmission is of particular importance in analyzing COVID-19 as it is one of the key features of the disease that makes controlling the outbreak difficult. Then, in our model, after the incubation period, at disease onset, members of *P* move to the infected class *I*. We note that another challenge with COVID-19 is that many patients will develop mild symptoms or none at all, yet being infectious. It is thus customary to collect these individuals in a separate compartment of asymptomatic individuals [18, 19]. Nevertheless, this distinction is not needed in our model as we will explain later in this section. Finally, patients transit to the removed compartment *R* by either recovery or by isolation after testing positive for COVID-19.

The above considerations are formulated in the following system of ordinary differential equations

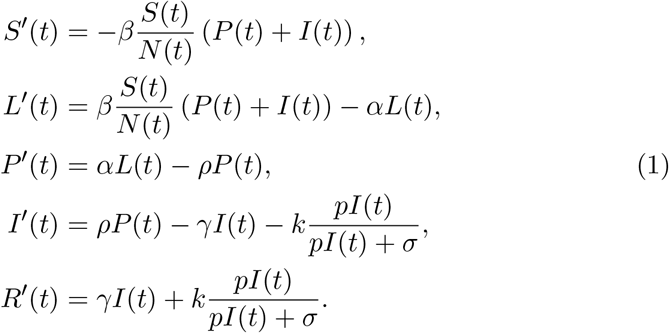

The disease transmission rate is denoted by the parameter *β*, the *incubation period* is *α*^−1^ +*ρ*^−1^, which is the sum of the duration of the latent period and the *pre-symptomatic* period, and, finally, *γ*^−1^ stands for the symptomatic infectious period. The transmission diagram of (1) is depicted on Fig. 1.

**Figure 1:**
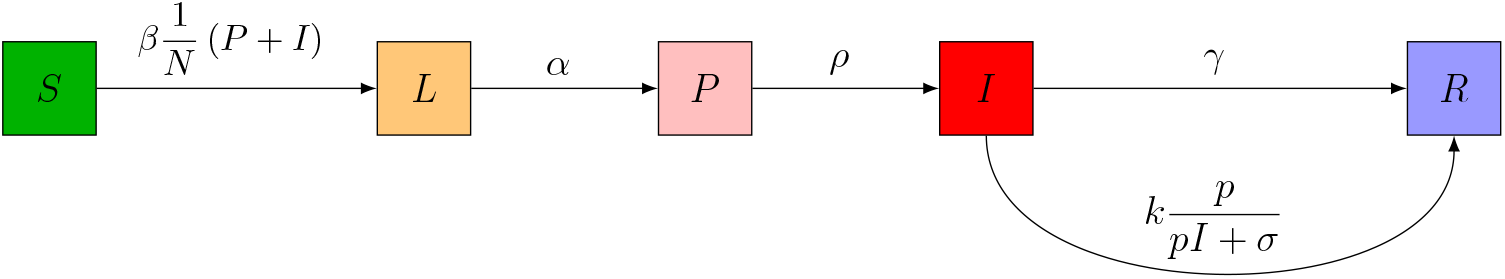
The transmission diagram of the *SLPIR* model (1). Arrows represent the transition rates between the compartments.

The *force of infection* is the rate associated with the outward flow from *S* to *L*, namely,

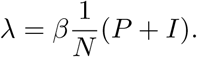

The indicator symptom-based testing is represented by the term

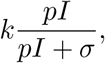

where *k* gives the number of tests done per unit time also referred to as the *testing rate*, the probability *p* describes how likely is that a member of compartment *I* displays the chosen indicator symptom. Note that this probability removes the need for an asymptomatic/mild compartment as it is straightforward to adjust *p* to account for all COVID-19 patients. The final term *σ* (possibly time-dependent) represents those individuals who are not infected by COVID-19, yet they show the very same symptom we base our testing upon. In this chapter, we refer to *σ* as the *secondary symptom pool*, whereas, the *primary symptom pool* Σ is composed of all members (with or without COVID-19 infection) of the population displaying the indicator symptom at a given time, that is

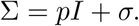

The testing rate *k* has a natural upper bound, namely,

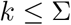

as we solely test patients displaying the indicator symptom. By reformulating the testing term as

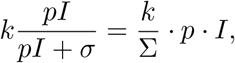

it is interpreted as the removal of the 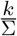 fraction of COVID-19 patients displaying the indicator symptom.

The rate of the testing-induced outward flow from *I* to *R* is referred to as the *force of testing* given by

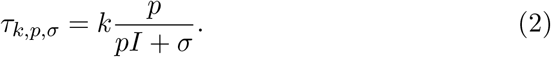

Finally, we introduce the *positivity rate* of testing as

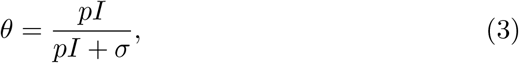

that may serve as a real-time indicator of the severity of an ongoing epidemic, and the adequateness of the testing rate.

Note that (1) is, in part, simpler than many other variants that have been used to assess the spread of COVID-19 as the infectious and latent compartments are not split into multiple stages [16, 18, 19, 22, 23]. However, these additional classes carry little significance for the testing strategies and to the analysis presented in this chapter. Hence, we chose to use this less complicated structure so that the emphasis is put on the testing itself.

We have parametrized (1) following [14]. From the infectivity profile of COVID-19 [3, 4, 5], we can see that most transmissions occur between 3 days prior to and 4 days after symptom onset, with the pre-symptomatic infection fraction being 43.7%. Thus, it is a reasonable approximation to set the pre-symptomatic period *ρ*^−1^ as 3 days, and the symptomatic infectious period *γ*^−1^ as 4 days, with the same infectiousness *β* during this period. The estimated mean incubation period of COVID-19 is 5.5 days [24], thus, the latent period *α*^−1^ is taken as 2.5 days, see Table 2. The choice of the transmission rate *β* is discussed in Sect. 2.1 and the testing parameters *k, p, σ* are varied throughout the analysis.

**Table 2:** Parameters of the *SLPIR* model

| Parameter | Notation | Value |
| --- | --- | --- |
| Transmission rate | $\beta$ | Sect. 2.1. |
| Latent period | $\alpha^{-1}$ | 2.5 days |
| Pre-symptomatic (infectious) period | $\rho^{-1}$ | 3 days |
| Infectious period | $\gamma^{-1}$ | 4 days |
| Testing rate | $k$ | varies |
| Secondary symptom pool | $\sigma$ | varies |
| Probability of symptom amongst COVID-19 patients | $p$ | varies |

### 2.1 Choosing the transmission rate *β*

Now, we concentrate on establishing the relationship between the transmission rate *β* in (1) and the basic reproduction number ℛ_0_ of the epidemic. We shall follow the terminology and techniques of [25] to compute the Next Generation Matrix (NGM) and the ℛ_0_ as its spectral radius.

First, let us consider the infectious subsystem of (1), namely, equations describing *L*(*t*), *P* (*t*), and *I*(*t*). Linearizing this subsystem w.r.t. the disease free equilibrium yields the linearized infectious subsystem

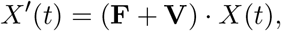

where the matrices **F** and **V** are referred to as the *transmission part* and *transitional part*, respectively; the state is described by

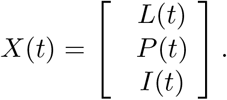

The transmission matrix **F** has the form

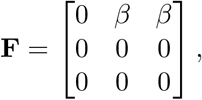

and the transitional matrix **V** is, clearly, written as

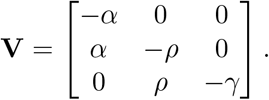

The basic reproduction number *R*_0_ is then obtained by computing the spectral radius of −**FV**^−1^ that is

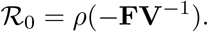

Therefore, as

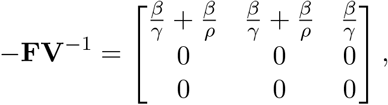

it follows that

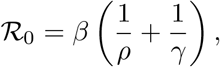

providing a scheme for computing *β*. We list the corresponding transmission rates for the sample values of *R*_0_ used for illustrations in Table 3.

**Table 3:** The basic reproduction number ℛ_0_ and the corresponding transmission rate *β*

|  |  |  |  |  |
| --- | --- | --- | --- | --- |
| $\mathcal{R}_0$ | 2.2 | 1.8 | 1.3 | 1.1 |
| $\beta$ | 0.338 | 0.277 | 0.2 | 0.169 |

The basic reproduction number ℛ_0_ is descriptive for the epidemic at the very beginning of an outbreak and in absence of control measures. For simplicity, we use the phrase basic reproduction number even if social distancing is in place, and by control measure in this chapter we mean the testing, the absence of which is modeled by *k* = 0. Similar key characteristics are the *control reproduction number* ℛ_*c*_ and the *effective reproduction number* ℛ_*t*_. The former describes the epidemic incorporating the effect of interventions, in our case indicator symptom-based testing, but still at the beginning of the outbreak. In contrast, the latter is suitable to measure the spread of the disease as the epidemic is progressing. The corresponding formulae may be obtained via analogous computations to those above as

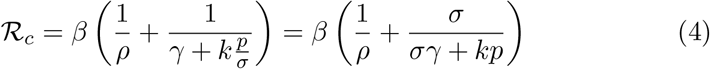

and

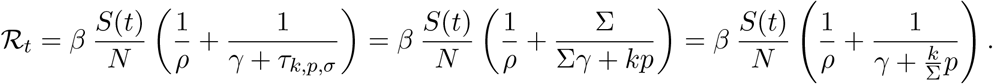

As the testing rate *k* is bound by the size of the primary symptom pool Σ, it is apparent that both of the above reproduction numbers satisfy

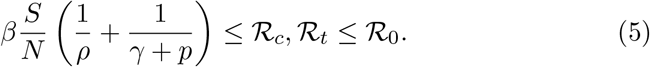

## 3 Dependence of key epidemic quantities on the testing strategy

This section analyzes the symptom-based testing strategy with emphasis on how the force of testing and the effective reproduction number are affected by the particular choice of strategy. Repeatedly, we shall utilize the monotonicity of

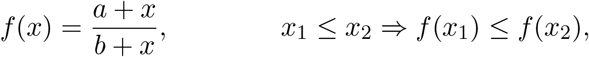

where 0 *< a* ≤ *b* and 0 ≤ *x*.

First, we summarize trivial monotonicity properties of the force of testing *τ*_*k,p,σ*_.

### Proposition 3.1.

*Given a fixed state of* (1), *the force of testing τ*_*k,p,σ*_ *is*

a. *monotonically increasing in k*,
b. *monotonically increasing in* 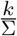.

In particular, as 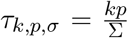, if 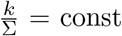, then *τ*_*k,p,σ*_ = const, *i.e*. the force of testing strongly correlates to what portion of the primary symptom pool is being tested.

As the epidemic is progressing, we may want to maintain the force of testing by increasing the testing rate *k* that is testing the same portion of individuals displaying the indicator symptom. Clearly, the required adjustment is linear w.r.t. the size of compartment *I*, thus, the given constant force of testing may be maintained as long as other logistical constraints make increasing the testing rate feasible.

The choice of the indicator symptom that serves as a basis for selecting patients for testing is clearly of importance. Different indicator symptoms typically have different associated probabilities and secondary symptom pools of non-equal sizes. Thus, it is natural to ask what (*p, σ*) pair is optimal.

### Proposition 3.2.

*The force of testing τ*_*k,p,σ*_ *is monotonically decreasing in* 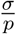.

*Proof*. Clearly,

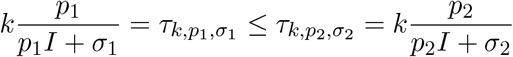

is equivalent to

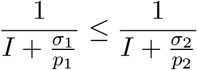

that, in turn, simplifies to

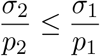

yielding the required result. □

As we have seen, keeping the fraction 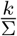 constant results in constant force of testing *τ*_*k,p,σ*_. The authorities might obtain some data on the size of the primary symptom pool Σ during an outbreak and use this information for adjusting *k* on-the-go. When planning for a second wave, historical data on the size of the secondary symptom pool *σ* may give information on the required level of preparedness. Namely, if we know that *σ* has now a different size compared to the former outbreak, *e.g*. due to a seasonal variation, we may utilize the size difference of the secondary symptom pools as a guidance for the need for testing capacities as follows.

### Proposition 3.3.

*Given a fixed state of* (1), *consider two secondary symptom pools*, 0 ≤ *σ*_1_ ≤ *σ*_2_ *for the same indicator symptom that appears amongst members of the compartment I with probability p. Let k*_1_ *and k*_2_ *be two testing rates corresponding to the testing strategies for σ*_1_ *and σ*_2_, *respectively. Then*,

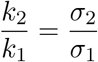

*implies*

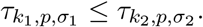

*Proof*. As the state is fixed, the two strategies having equal effect corresponds to the equality

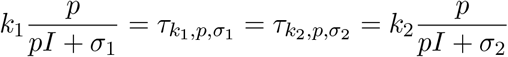

that simplifies to

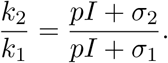

Then, using the aforementioned monotonicity of *f* (*x*), we obtain

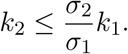

Finally, the monotonicity of *τ*_*k,p,σ*_ in *k* completes the proof. □

Recall, that the force of testing *τ*_*k,p,σ*_ explicitly appears in the formula for the effective reproduction number ℛ_*t*_ as

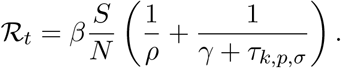

Accordingly, ℛ_*t*_ may be kept decreasing by varying *k* as discussed in the first half of this section that is by keeping *τ*_*k,p,σ*_ constant or increasing. However, in practice, increasing *k* may eventually become infeasible. At that point, the force of testing will decrease, hence, ℛ_*t*_ may increase temporarily, within the bounds given in (5), despite the constantly decreasing number of susceptible individuals *S*(*t*).

A reasonable goal for the authorities is to keep ℛ_*t*_ close to a designated value, ideally close to 1 to suppress the epidemic. Running estimates of the actual ℛ_*t*_ might be obtained [19, 27, 28], hence, we investigate if, by an increase of the testing rate, we can alter ℛ_*t*_ as desired.

### Proposition 3.4.

*Let* 0 ≤ *k*_1_ ≤ *k*_2_ *be two testing rates. Consider an epidemic described by* (1) *with daily testing rate k*_1_, *and the associated effective reproduction number* ℛ_*t*_(*k*) *as a function of k*.

*Then, the ratio of the effective reproduction numbers corresponding to altering the testing rate from k*_1_ *to k*_2_

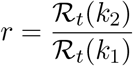

*satisfies the following inequality*

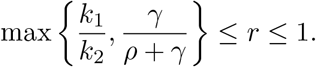

*Proof*. The right bound is trivial as ℛ_*t*_ is monotonic in *k*. Now, observe that

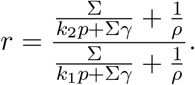

Then,

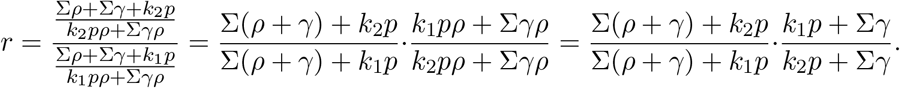

The first term is ≥ 1, thus,

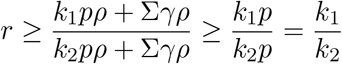

using the monotonicity of *f* (*x*) noted at the beginning of this section.

Now, consider reordering the product as

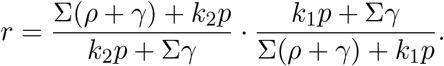

Again, the first term is ≥ 1, therefore,

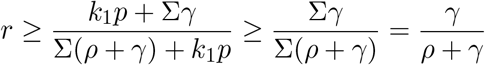

holds using, again, the monotonicity of *f* (*x*).

Combining the two inequalities above completes the proof. □

The implications of Prop. 3.4 on goals for the testing strategy are rather important as they point out some hard limitations. Clearly, as

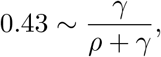

no matter our testing capacity or indicator symptom, we may not suppress the epidemic any further. As an example, if our current estimates for ℛ_*t*_ are above 2.4, then we cannot expect the pure indicator symptom-based testing strategy (without contact-tracing) to be able to suppress the epidemic as 2.4 · 0.43 ∼ 1.03. Additionally, as the indicator symptom limits our testing rate to *k* ≤ Σ = *pI* + *σ*, we obtain another hard constraint, namely,

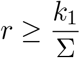

that is the ratio describing what proportion of the primary symptom pool is being tested directly limits the factor which the effective reproduction number may be decreased with via larger testing rates. Finally, we note that reordering the inequality yields 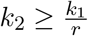 as a lower requirement for the required testing rate – given that the reduction by factor *r* is achievable.

We have discussed from various aspects that increasing the testing rate *k* decreases the effective reproduction number ℛ_*t*_ that is it has a positive effect on the severity of the epidemic. Nevertheless, this positive effect is gradually decreasing as described by the following Proposition.

### Proposition 3.5.

*Consider the logarithmic derivative of* ℛ_*t*_ *w.r.t. the testing rate k that is*

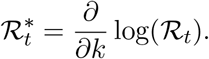

*Then*, 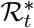 *is negative and monotonically increasing in k*.

*Proof*. Clearly,

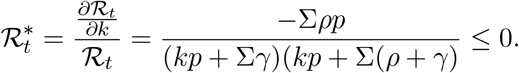

Then,

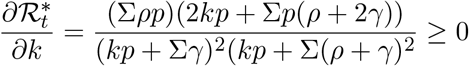

completes the proof. □

This logarithmic derivative is a measure of the relative change in ℛ_*t*_ w.r.t. the testing rate *k*. Prop. 3.5 states that the relative change is decreasing in absolute value as *k* increases.

## 4 Numerical simulations

This section presents the results from several numerical simulations demonstrating the impact of the key parameters of the epidemic model (1). All simulations were executed with a sample population of size 10, 000, 000 with initial conditions placing 1000 individuals into the class *L* and the rest into *S*.

First, Sect. 4.1 presents the numerical analysis of the control reproduction number ℛ_*c*_. Then, we investigate the connection between the progress of an outbreak and the positivity rate of testing in Sect. 4.2. We study the implications of maintaining a constant force of testing *τ*_*p,k,σ*_ in Sect. 4.3. The significance of the seasonality of the secondary symptom pool *σ* is analyzed in Sect. 4.4. Finally, in Sect. 4.5, we assess how an increased testing rate may delay the progress of COVID-19.

### 4.1 The effect of testing on the control reproduction number ℛ_*c*_

The control reproduction number ℛ_*c*_, given in (4), describes the initial progress of the epidemic at its very beginning. Fig. 2 demonstrates what effect of indicator symptom-based testing has on ℛ_*c*_ for various values of ℛ_0_ and *σ*.

**Figure 2:**
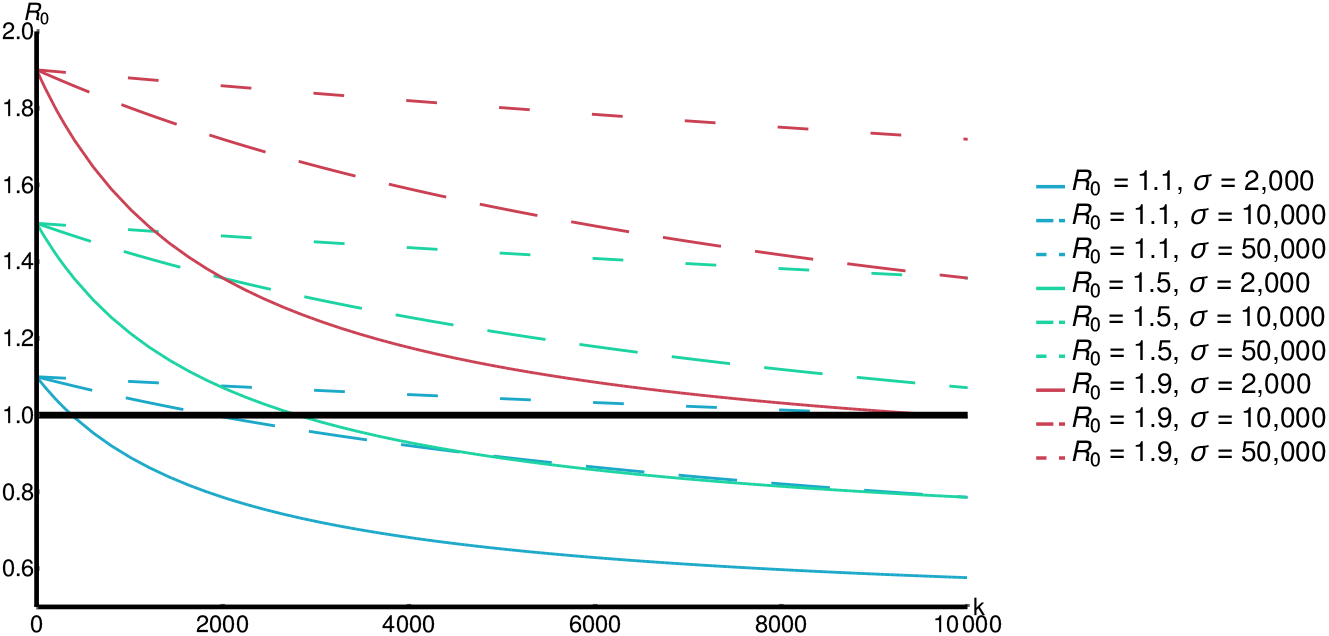
The effect of indicator symptom-based testing on ℛ_*c*_ for *p* = 0.25.

Clearly, larger maximal testing rate *k* results in lower ℛ_*c*_. The size of the secondary symptom pool *σ* apparently greatly affects the decrease we may achieve by larger *k*.

### 4.2 The progress of an outbreak and the positivity rate *θ*

Recall that the positivity rate *θ*, see (3), is a key feature of the testing strategy that may be readily observed during an outbreak. If the efforts aimed at suppressing COVID-19 are not successful, the rate *θ* will increase as the term *pI*(*t*) will eventually dominate the secondary symptom pool *σ*. Fig. 3 demonstrates that the changes in *θ* are in close connection with the dynamics of *I*(*t*). This relationship between *θ* and *I*(*t*) carries a certain benefit for the authorities as the increase of the positivity rate precedes that of the epidemic curve, hence, it may serve as a primary indicator for the progress of an epidemic.

**Figure 3:**
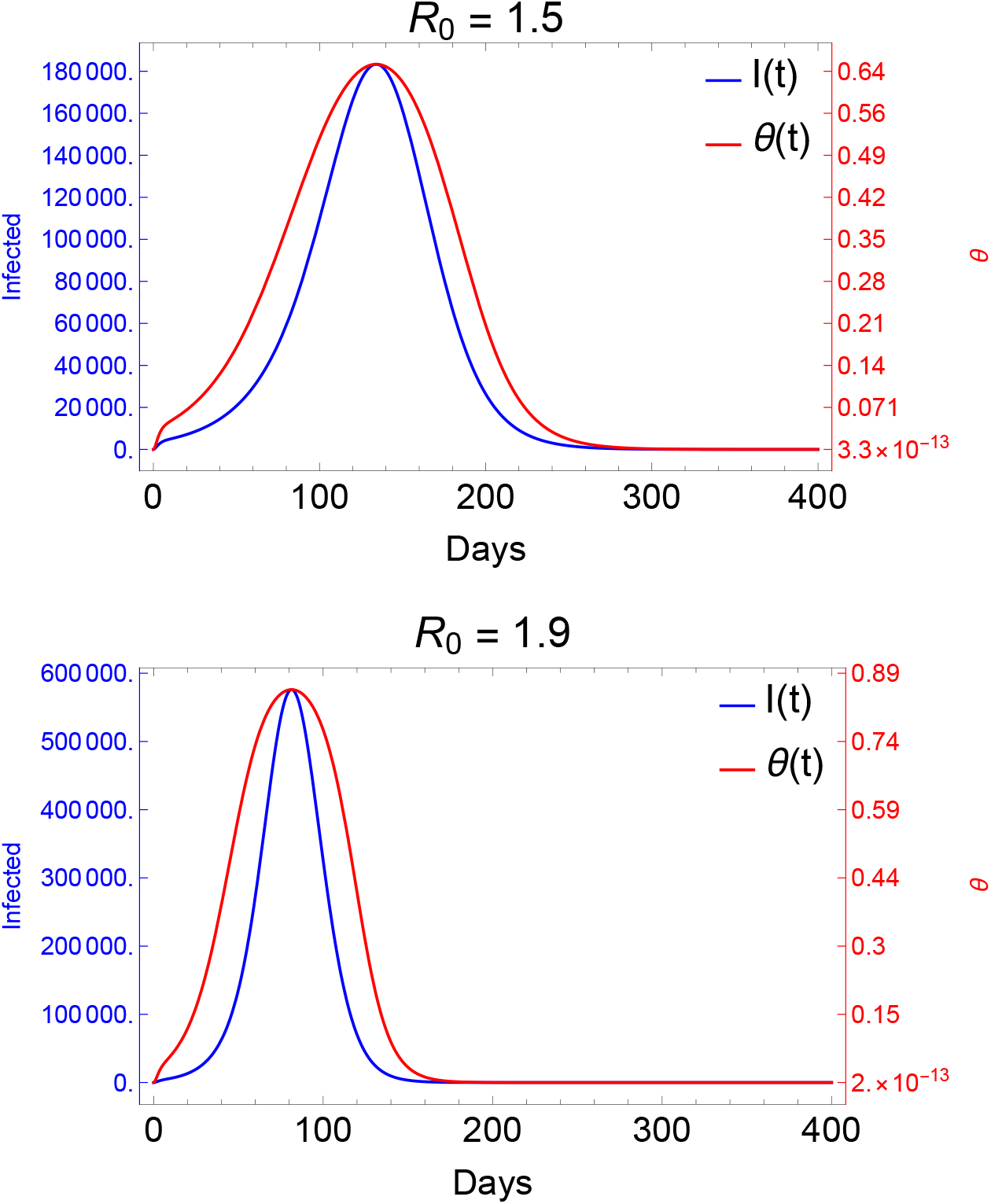
Evolution of the positivity rate during outbreaks of different magnitudes. The prevalence of the indicator symptom is *p* = 0.1 with a maximal testing capacity *k* = 10,000 and secondary symptom pool *σ* = 10,000.

### 4.3 Implications of constant force of testing *τ*_*p,k,σ*_

As we have discussed in Sect. 3, a constant force of testing *τ*_*p,k,σ*_ is achieved by testing a fixed portion of the primary symptom pool Σ, *i.e*. 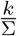 is constant. For an ongoing epidemic this results in a constant increase in the required daily testing rate *k*. We have analyzed the maximal required testing capacity w.r.t. COVID-19 patients in Fig. 4.

**Figure 4:**
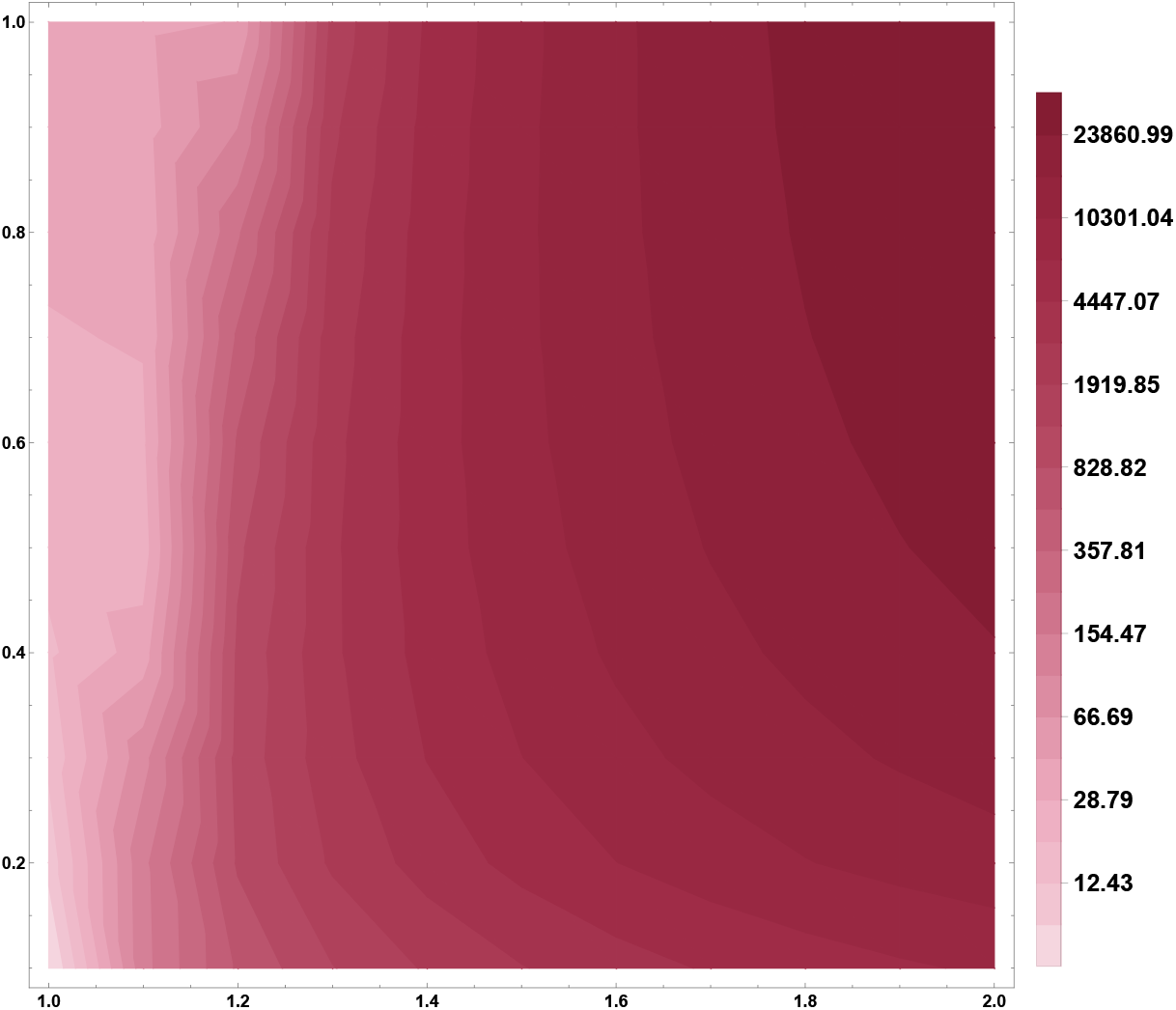
Required testing capacity to maintain a constant force of testing *τ*_*p,k,σ*_. The vertical axis describes the desired portion for testing the primary symptom pool and the horizontal axis represents the underlying basic reproduction number ℛ_0_. The prevalence of the indicator symptom is set to *p* = 0.1.

Note that for constant *τ*_*p,k,σ*_, the system (1) is independent of the secondary symptom pool *σ*, thus, this requirement must be adjusted based on historical data on the size of *σ* to obtain the total maximal required capacity.

### 4.4 Seasonality of the secondary symptom pool *σ*

Now, let us investigate the epidemic curves in case of a periodically varying secondary symptom pool. To that end, we employ a commonly used seasonality function

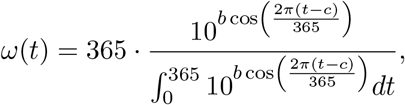

with *b* = 0.5 and consider *σ* = *σ*_avg_ · *ω*(*t*). The parameter *c* is used to model shift in the seasonality, *i.e*. to analyze the differences between an outbreak starting at minimal or maximal secondary symptom pools. The function *ω*(*t*) is displayed on Fig. 5 for the case of minimal secondary symptom pool at time *t* = 0 that is for a shift *c* = 183.

**Figure 5:**
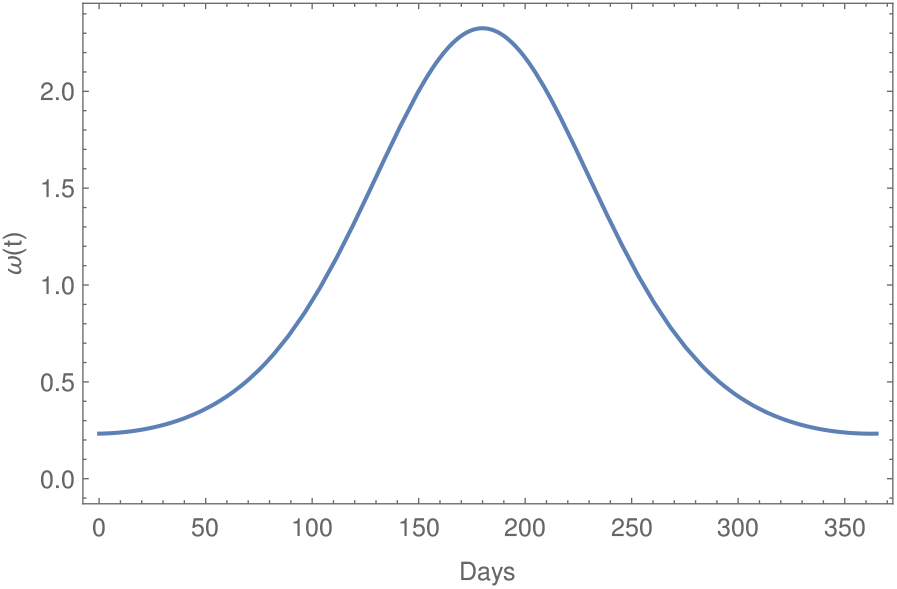
The seasonality function *ω*(*t*) with *c* = 183. This corresponds to minimal secondary symptom pool at the beginning of an outbreak.

Fig. 6 demonstrates the effect of having seasonality in *σ* and the COVID-19 outbreak beginning around the minimal size of the secondary symptom pool. This comparison shows that we may expect a slight, but notable, delay in this scenario compared to the non-seasonal setting.

**Figure 6:**
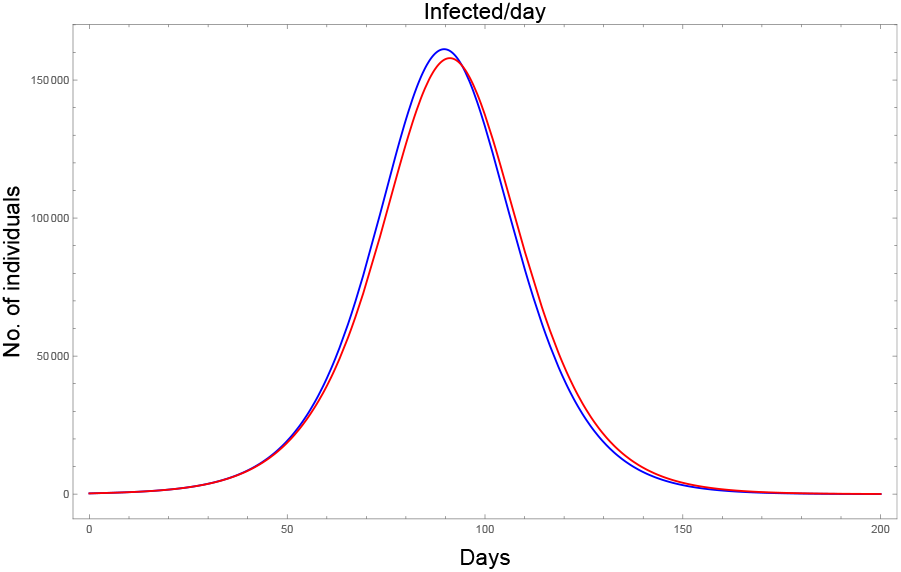
The impact of seasonal *σ* with minimal size at the beginning of the outbreak. ℛ_0_ = 1.9, *p* = 0.1, *k* = 10,000, *σ* = 10,000. The blue curve corresponds to assuming a constant (average) secondary symptom pool, whilst, the red curve depicts the effect of seasonality.

A similar shift in the opposite direction takes place if we consider the beginning of the outbreak to coincide with the maximal state of *σ*, see Fig. 7.

**Figure 7:**
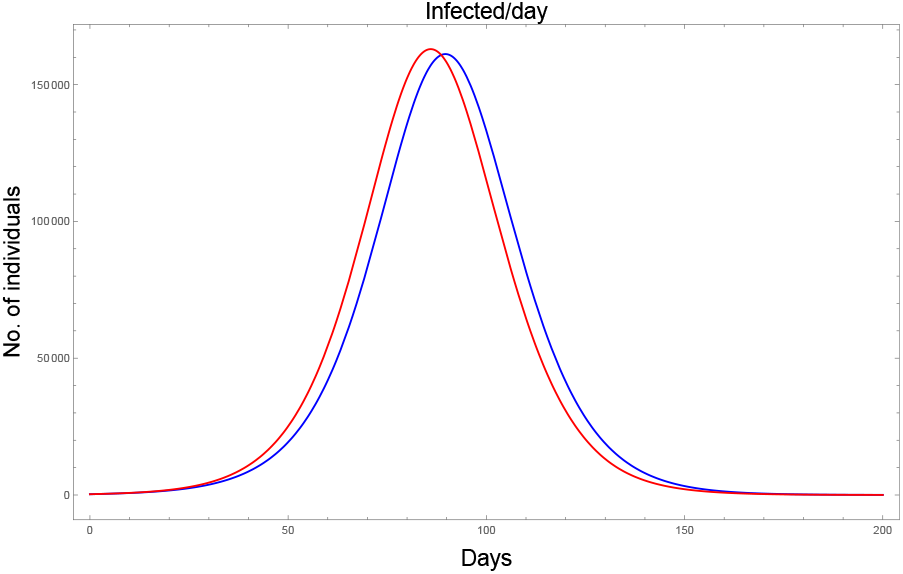
The impact of seasonal *σ* with maximal size at the beginning of the outbreak. ℛ_0_ = 1.9, *p* = 0.1, *k* = 10,000, *σ* = 10,000. The blue curve corresponds to assuming a constant (average) secondary symptom pool, whilst, the red curve depicts the effect of seasonality.

### 4.5 The effect of varying the testing rate *k*

Increasing the testing rate *k* has a beneficial effect. We demonstrate this via transitional plots on Fig. 8. Note that a larger maximal *k* both delays in time and decreases in size the peak of the epidemic.

**Figure 8:**
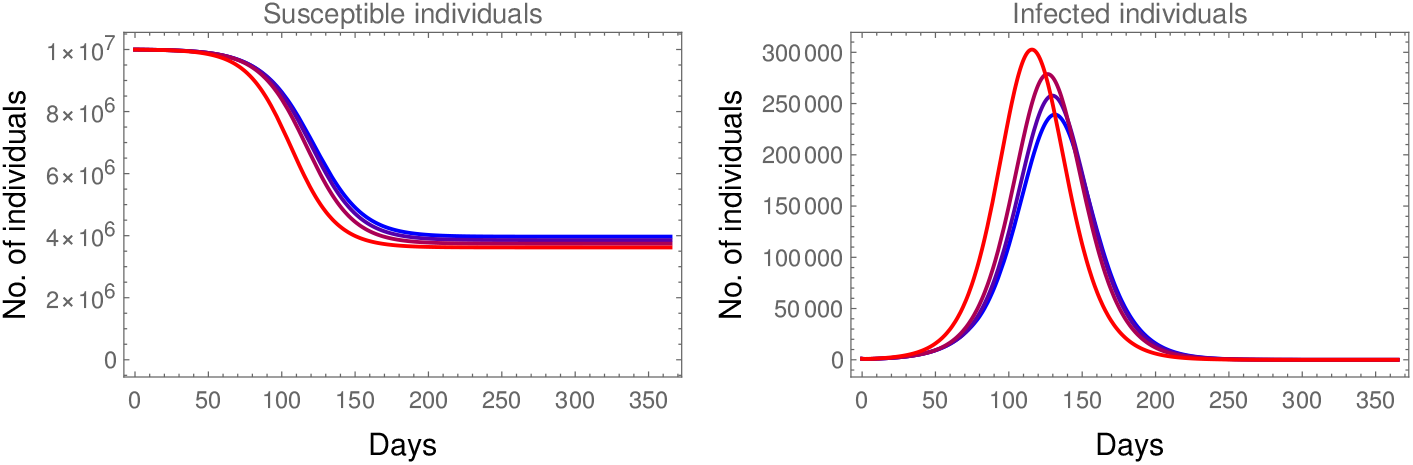
The impact of increasing the testing rate from 1,000 (red) to 10,000 (blue) using parameters ℛ_0_ = 1.6, *p* = 0.1, 1, 000 ≤ *k* ≤ 10,000, *σ* = 10,000.

## 5 Conclusions

We have investigated the effects of indicator symptom-based testing on COVID-19. The benefits of increasing the testing rate *k* are demonstrated, suggesting that, as long as other logistical constraints allow, the authorities should aim to keep it as high as possible. The choice of the indicator symptom is of importance. We have shown that not just its prevalence *p* should be taken into account but the size and seasonality of the associated secondary symptom pool *σ* as well. Note that the analysis in this chapter did not directly consider contact/transmission-reducing nonpharmaceutical interventions (NPIs), *i.e*. curfew, closures of schools, wearing of masks, etc. Naturally, these interventions would affect not just the spread of COVID-19, but of other diseases, hence, potentially decreasing the secondary symptom pool *σ* as well. Such NPIs may be fitted into the presented framework by varying the basic reproduction number ℛ_0_ and *σ*, as seen in Sect. 4.1.

The quality of tests was not considered. The false negativity rate could be easily modeled by a reduction factor in *k*. Handling the false positivity rate is more involved as susceptible individuals (susceptible to COVID-19, but still displaying the indicator symptom, *i.e*. members of *σ*) may be temporarily removed from the infectious chain just to reappear later, after a precautionary quarantine. However, rRT-PCR-tests have very high specificity, hence, false positives are rare.

We have modeled the transmission of COVID-19 using identical rates for the presymptomatic *P* and symptomatic *I* classes. This choice is influenced by the current understanding that according to the inferred infectivity profiles, the transmissibility prior to and after the onset of symptoms is of similar magnitude, and the ratio of presymptomatic transmissions is almost 50% [3, 4, 5]. Nevertheless, using different rates for the two compartments would not alter the computations heavily.

It is clear from the numerical simulations that indicator symptom-based testing, alone, cannot prevent an outbreak. It has a modest effect in delaying and slowing down the epidemic. Thus, symptom based testing alone may have clinical importance by providing guidance about how to treat a given patient, but its impact as epidemic mitigation is negligible. Therefore, in practice, authorities should opt to perform agile contact-tracing based on positive COVID-19 tests. The effect of this additional intervention is not included in our analysis. Nevertheless, it is safe to claim that the addition of contact-tracing would considerably increase the benefits of any testing strategy, in particular, some individuals would get removed from the presymptomatic compartment *P* and the latent compartment *L* as well via additional testing or general quarantine for contacts of COVID-19 patients with positive test result.

In summary, testing and isolation of cases is a key tool in combating the pandemic. However, symptom-based testing alone is not sufficient to control COVID-19. To significantly ease the disease burden on the society, it must be used in combination with other measures.

## Data Availability

All data used is properly cited.

## Acknowledgements

This work was done in the framework of the Hungarian National Development, Research, and Innovation (NKFIH) Fund 2020-2.1.1-ED-2020-00003 and of the grants TUDFO/47138-1/2019-ITM and EFOP-3.6.2-16-2017-00015. Some authors were also supported by NKFIH KKP 129877 (J.K.), NKFIH FK 124016 (T.T.), János Bolyai Research Scholarship of the Hungarian Academy of Sciences (F.B.).

